# Effect of removing race correction factor in glomerular filtration rate estimation on predicting acute kidney injury after percutaneous coronary intervention

**DOI:** 10.1101/2022.01.18.22269155

**Authors:** Chenxi Huang, Karthik Murugiah, Xumin Li, Frederick A. Masoudi, John C. Messenger, Bobak J. Mortazavi, Harlan M Krumholz

## Abstract

**Background:** Race is a social and not a biological construct. Major societies have recommended removing a race correction factor when estimating glomerular filtration rate (eGFR). eGFR is a strong predictor for acute kidney injury (AKI) after percutaneous coronary intervention (PCI). We assessed the effect of removal of the race correction factor in eGFR calculation on the predictive ability of contemporary models for AKI after PCI.

**Methods:** We used data from the American College of Cardiology’s National Cardiovascular Data Registry (NCDR) CathPCI registry to assess the effect of removing a race correction in eGFR calculation on two previously published AKI models developed using NCDR data – a logistic regression-based model and a gradient boosting machine learning (ML) model. We first assessed the calibration and discrimination of these models with and without race correction and subsequently included race as an independent predictor in a model without race-corrected eGFR. We assessed model performance overall, and stratified by Black and non-Black subgroups. The models were trained on the same cohort used to develop the NCDR models and validated with a contemporaneous validation data set.

**Results:** We included 947,091 PCI procedures in 915,223 patients (mean age 64.8 years; 7.9% were Black and 32.8% women) with an AKI rate of 7.4%. In the NCDR model, inclusion of race correction in eGFR significantly underestimated AKI risk among Black patients (predicted 7.6% vs observed 10.2%) while slightly overestimating risk among non-Black patients (predicted 7.4% vs observed 7.1%). Removing the race correction partially corrected the underestimation among Black patients (predicted 8.2%). Including race as an independent predictor and introducing interaction terms further reduced the underestimation (predicted 10.1%). The receiver-operating-characteristic curve (AUC) was similar among these models, but was consistently lower in Black patients. Compared with the logistic model, the ML model had better calibration in Black patients (with race correction predicted 8.6% and without 8.7%) but still underestimated AKI risk. With race included as a predictor, the ML model achieved similarly good calibration in Black patients (predicted 10.1%). The AUCs of ML models were better than the logistic models but did not differ based on the inclusion of the race correction.

**Conclusion:** Removing the race correction in eGFR calculation has a positive effect of reducing the underestimation of the risk of AKI following PCI for Black patients. However, despite the reduction in underestimation, Black patients remain at elevated risk for AKI compared to non-Black patients. There is a need to better capture the determinants of this higher AKI risk through richer data and advanced modeling techniques.

## INTRODUCTION

The inclusion of race correction to estimate the glomerular filtration rate (eGFR) is controversial.(1) The two currently used creatinine-based eGFR equations, the Modification of Diet in Renal Disease (MDRD) and Chronic Kidney Disease Epidemiology Collaboration (CKD-EPI), use a correction factor for Black race.(2,3) In an acknowledgement that race is a social construct with only rare biological significance, the National Kidney Foundation (NKF) and the American Society of Nephrology (ASN) task force have recommended the removal of race correction factor in eGFR calculation(4) and new eGFR equations are being developed excluding the race variable.(5)

A reduced eGFR is one of the strongest predictors for acute kidney injury (AKI) after percutaneous coronary interventions (PCI) in the National Cardiovascular Data Registry (NCDR)-AKI model (6). Also, physicians routinely use the eGFR interchangeably with creatinine clearance to estimate the maximum contrast limit during PCI.(7,8) The removal of the race correction factor will change the risk estimates of contemporary models thus affecting calculated risk estimates informing clinical decisions. There is a need to reevaluate in detail the performance of the current models if the race correction factor is removed.

Accordingly, we sought to evaluate the impact of removal of race in eGFR calculation on the accuracy of risk prediction for AKI after PCI with data from the American College of Cardiology’s National Cardiovascular Data Registry (NCDR) CathPCI Registry. We compared the calibration and discrimination of the current logistic regression based NCDR AKI model (NCDR model) with and without a race correction factor. We then similarly assessed the impact of inclusion and exclusion of race in eGFR calculation on our previously published Extreme Gradient Boosting (XGboost) based machine learning risk model (ML model).(9) Finally, we evaluated model performance when removing race from eGFR calculation but including it as an independent predictor.

## METHODS

### Study population

The NCDR CathPCI registry has been described previously. The registry contains data on PCI from >1000 sites across the United States. Abstractors collect data in a standard format and the registry is monitored by a comprehensive data quality program.

We used the same cohort that was used to develop the NCDR model(6) and the subsequent ML model(9), including all PCI between June 1, 2009 and June 30, 2011. We also applied the same exclusion criteria, including subsequent procedures during a single hospitalization, procedures with same-day discharge, missing serum creatinine before or after PCI, and those for patients already on dialysis at the time of PCI. The models were trained using the remaining 947,091 procedures. A more contemporary cohort of procedures between July 1, 2011 and June 30, 2017 were used as an independent validation cohort to evaluate accuracy of the risk predictions. The study was approved by the Institutional Review Board at Yale University.

### Study outcome and variables

The primary outcome, post-PCI AKI, was defined as a change in post-procedural creatinine larger than 0.3 mg/dL or 50% from before the procedure, according to the Acute Kidney Injury Network (AKIN) and the definition used in developing the NCDR model.(10) We calculated the eGFR according to the MDRD equation, the same formula used in estimating the eGFR as a predictor included in the NCDR model. A race correction factor of 1.212 was used for African Americans in this equation and the race variable from the NCDR data was used for the calculation.

### Statistical analysis

The development and predictors of the NCDR model and the ML model have been previously described in detail.(6,9) For this study, we trained models on a random 70% of the cohort (training set) and evaluated the performance of the models on both the remaining 30% of the cohort (test set) and the validation cohort. Models were re-calibrated on the validation cohort by Platt’s Scaling. For both the NCDR and ML model, eGFR is a predictor and a series of models were trained by changing how eGFR was calculated, including a model with race correction factor included in the eGFR calculation (Model 1), a model with the race correction factor excluded (Model 2) and a model including race as an additional and independent predictor (Model 3). For the NCDR model, we considered the inclusion of interaction terms between race and other predictors via likelihood ratio tests. The XGBoost based ML model is able to incorporate interaction terms without explicit specifications, so no additional model selection step was conducted.

The model performance was assessed for discrimination by the area under the receiver-operating-characteristic curve (AUC) and calibration by comparing the average predicted risks with observed risks (i.e., calibration-in-the-large) and visually assessing the calibration curves, which plot the observed risks versus predictions for deciles of predicted risks. The performance metrics were calculated in the entire cohort and in Black and non-Black patients respectively.

R (version 3.6.0) was used for all analyses.

## RESULTS

### Patient characteristics

The cohort included 947,091 PCI encounters with an average patient age of 64.8 years; 74,820 (7.9%) patients were Black and 311,013 (32.8%) were women. The observed AKI rate was 7.4%. The validation cohort included 3,063,853 procedures with an AKI rate of 7.5%. The patients included in the validation cohort had an average age of 65.3 years; 261,384 (8.5%) African Americans and 976,924 (31.9%) women.

### Impact of removal of race correction factor on the NCDR model

The results of comparing models using the logistic regression based NCDR model are shown in Table 1. Evaluated in the test set, the AUCs did not differ significantly for models with or without race correction factor in calculating the eGFR (Model 1 and 2), in the overall cohort population and in both Black and non-Black groups. The AUCs for both models were slightly worse in the Black group compared with the non-Black group (0.68 vs 0.72).

**Table 1.** Performance comparison of logistic regression models in internal test set.

|  | Calibration |  |  |  |  |  | Discrimination |  |  |
| --- | --- | --- | --- | --- | --- | --- | --- | --- | --- |
|  | Mean risk in overall population, % |  | Mean risk in Black patients, % |  | Mean risk in non-Black patients, % |  | AUC in overall population | AUC in Black patients | AUC in non-Black patients |
|  | Observed | Predicted | Observed | Predicted | Observed | Predicted |  |  |  |
| Model 1 | 7.4(7.3-7.5)* | 7.4(7.4-7.4) | 10.2(9.8-10.6) | 7.6(7.5-7.7) | 7.1(7.0-7.2) | 7.4(7.4-7.4) | 0.71(0.71-0.72) | 0.68(0.67-0.70) | 0.72(0.71-0.72) |
| Model 2 | 7.4(7.3-7.5) | 7.4(7.4-7.4) | 10.2(9.8-10.6) | 8.2(8.1-8.3) | 7.1(7.0-7.2) | 7.3(7.3-7.4) | 0.71(0.71-0.72) | 0.68(0.67-0.70) | 0.72(0.71-0.72) |
| Model 3 | 7.4(7.3-7.5) | 7.4(7.4-7.4) | 10.2(9.8-10.6) | 10.1(10.0-10.2) | 7.1(7.0-7.2) | 7.2(7.1-7.2) | 0.72(0.71-0.72) | 0.68(0.67-0.70) | 0.72(0.71-0.72) |

\*Results are shown in mean (95% confidence interval).
Model 1: Logistic regression model with race factor included in the eGFR calculation. Model 2: Model with race factor removed from eGFR calculation. Model 3: Model with interactions between race and age, prior cardiogenic shock, cardiac arrest, baseline eGFR, anemia, CAD presentation, diabetes and heart failure within 2 weeks.
AUC: area under the receiver operating characteristic curve; eGFR: estimated glomerular filtration rate; CAD: coronary artery disease.

Model 1 showed good calibration in the overall population with slight overestimation in the non-Black group (predicted vs observed 7.4% vs 7.1%), but significant underestimation of risk in the Black group (predicted vs observed 7.6% vs 10.2%). In Model 2 the underestimation in the Black group was slightly lower than Model 1(predicted vs observed 8.2% vs 10.2%).

Tests for interaction terms of race as an independent predictor with other predictors resulted in a model (Model 3) including interaction terms of race with age, prior cardiogenic shock, prior cardiac arrest, baseline kidney function, anemia, presence of coronary artery disease, diabetes, and heart failure within the prior 2 weeks. This model further reduced the underestimation of risks in Black patients (predicted vs observed 10.1% vs 10.2%) while the AUC was similar to Model 1 and Model 2. The overestimation among non-Black group was also reduced (predicted vs observed 7.2% vs 7.1%). Similar results were found in evaluating the models in the validation cohort (Supplemental Table S1).

The calibration plots for Model 1 visually showed underestimation of risks in the Black group, especially in the higher risk strata (Figure 1). Model 2 showed reduction of underestimation for Black patients and Model 3 showed further improvement with good calibration across all risk groups. The non-Black group had good calibration with or without race correction factor in the eGFR calculation.

**Figure 1.**
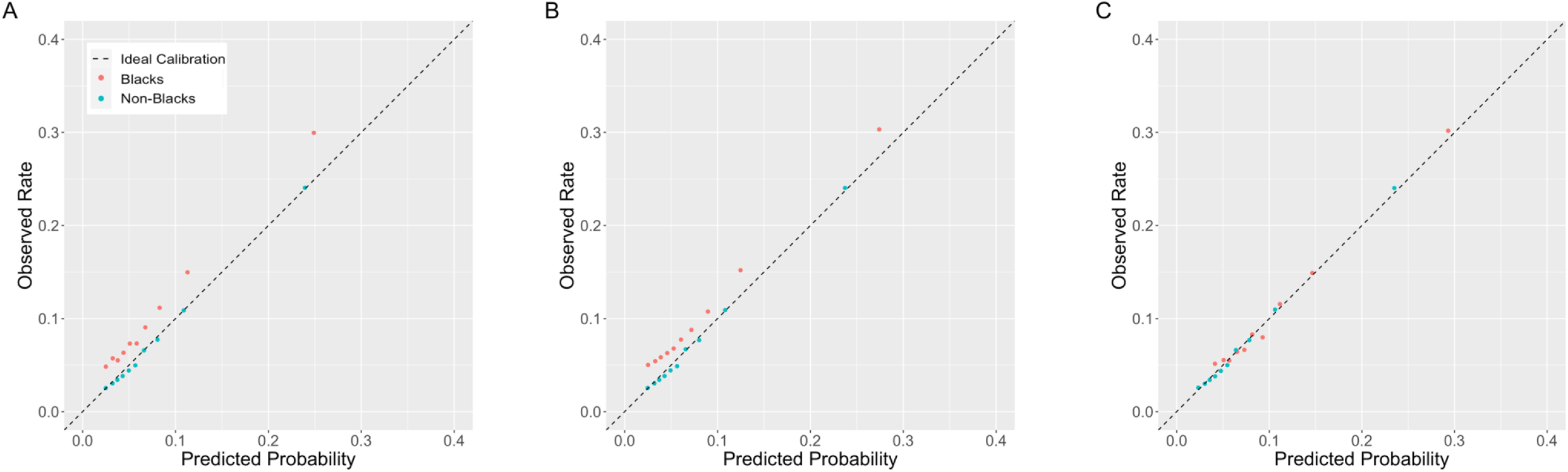
Calibration plots of logistic regression models for Black and non-Black groups for (A) model with race factor included in the eGFR calculation, (B) model with race factor removed in the eGFR calculation, and (C) model with interaction terms of race and other predictors. The plots show observed risk of acute kidney injury (AKI) versus mean predictions in deciles of predicted AKI risks.

### Impact of removal of race correction factor on machine learning (ML) model

Compared with the NCDR models, the ML models achieved higher AUCs, but similar to the logistic regression models they did not differ across models with or without race correction factor in eGFR calculation (Table 2).

**Table 2.** Performance comparison of machine learning models in internal test set.

|  | Calibration |  |  |  |  |  | Discrimination |  |  |
| --- | --- | --- | --- | --- | --- | --- | --- | --- | --- |
|  | Mean risk in overall population, % |  | Mean risk in Black patients, % |  | Mean risk in non-Black patients, % |  | AUC in overall population | AUC in Black patients | AUC in non-Black patients |
|  | Observed | Predicted | Observed | Predicted | Observed | Predicted |  |  |  |
| Model 1 | 7.4(7.3-7.5) | 7.4(7.4-7.4) | 10.2(9.8-10.6) | 8.6(8.5-8.7) | 7.1(7.0-7.2) | 7.3(7.3-7.3) | 0.75(0.75-0.76) | 0.71(0.70-0.73) | 0.76(0.75-0.76) |
| Model 2 | 7.4(7.3-7.5) | 7.4(7.4-7.4) | 10.2(9.8-10.6) | 8.7(8.6-8.8) | 7.1(7.0-7.2) | 7.3(7.2-7.3) | 0.75(0.75-0.76) | 0.72(0.70-0.73) | 0.76(0.75-0.76) |
| Model 3 | 7.4(7.3-7.5) | 7.4(7.4-7.4) | 10.2(9.8-10.6) | 10.1(9.9-10.2) | 7.1(7.0-7.2) | 7.2(7.3-7.5) | 0.75(0.75-0.76) | 0.72(0.71-0.73) | 0.76(0.75-0.76) |

\*Results are shown in mean (95% confidence interval).
Model 1: Machine learning model with race factor included in the eGFR calculation. Model 2: Model with race factor removed from eGFR calculation. Model 3: Model with race as an independent predictor.
AUC: area under the receiver operating characteristic curve; eGFR: estimated glomerular filtration rate; CAD: coronary artery disease.

Similar to the NCDR models, the ML Model 1 underestimated AKI risks for the Black group (predicted vs observed is 8.6% vs 10.2%) while ML Model 2 reduced the underestimation (predicted vs observed is 8.7% vs 10.2%). While underestimating risks in Black patients, the risk predictions of the ML models were closer to the observed risks than predictions of the logistic regression models, with or without the race correction factor. Including race as an additional predictor, the updated ML model (Model 3) achieved good agreement between the predicted and observed risks in Black patients (predicted vs observed is 10.1% vs 10.2%). Similar results were found in evaluating the models in the validation cohort (Supplemental Table S2).

Calibration plots showed better calibration across risk groups for the ML models compared to the logistic regression models (Figure 2). Removing the race correction factor in the ML model achieved better calibration, especially in the highest risk group. Adding race as a predictor, the ML model had good calibration in all risk groups. The non-Black group by the ML models achieved good calibration regardless of whether race correction factor was included in eGFR.

**Figure 2.**
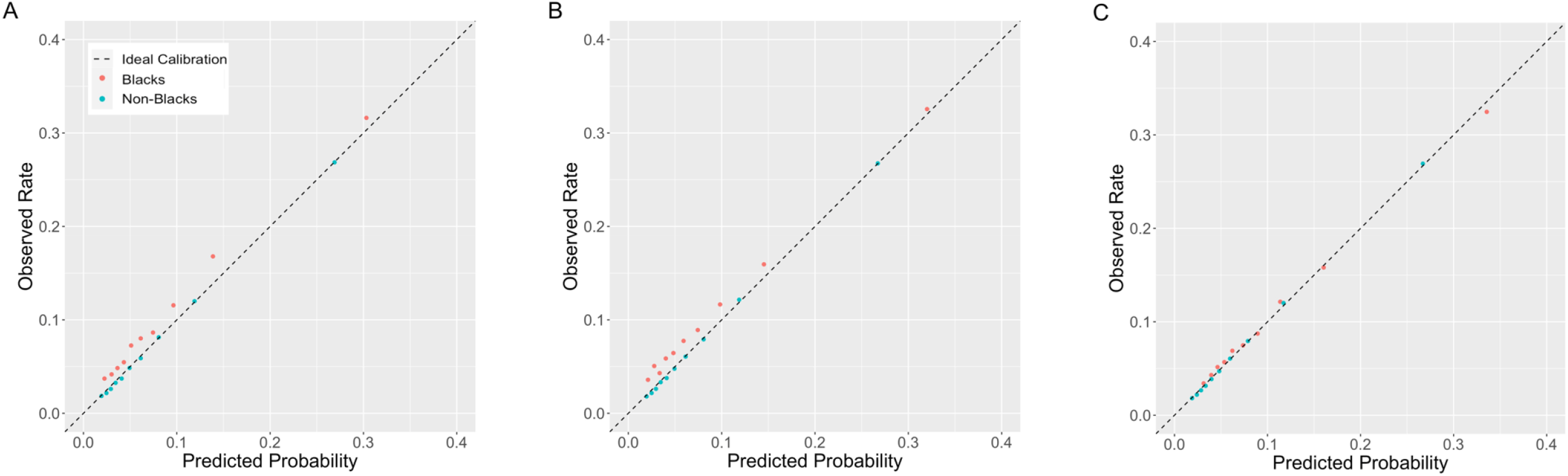
Calibration plots of machine learning models for Black and non-Black groups for (A) model with race factor included in the eGFR calculation, (B) model with race factor removed in the eGFR calculation, and (C) model with race as an independent predictor. The plots show observed risk of acute kidney injury (AKI) versus mean predictions in deciles of predicted AKI risks.

## DISCUSSION

In this national registry study, we found that the current inclusion of the race correction factor in eGFR calculation causes a systematic underestimation of the AKI risk following PCI among Black patients regardless of the model used. Removing the race correction factor leads to better model calibration for Black patients, who are at a higher overall risk for AKI than non-Black patients. Including race as a covariate and its interactions with other risk factors leads to significant further improvements in calibration.

The inclusion of race in the original MDRD and CKD-EPI equations was based on data which showed that among patients with similar measured GFR the serum creatinine levels were higher among adults who identified themselves as Black.(2,11) However, the inclusion of race in the eGFR equation has important shortcomings. It ignores the ancestral diversity among Black patients and patients with multiracial background and introduces bias due to the subjectivity in self-identification of race. The NKF and ASN task force recommends an immediate removal of the race correction factor from eGFR estimation.(4)

However, removal of the race factor and the resulting change in eGFR can potentially influence diagnostic and treatment decisions. The NKF and ASN task force in their deliberations reviewed a few common scenarios such as kidney disease screening and transplant referral, medication candidacy and dosing, and eligibility for clinical trials to understand the effect of removal of race correction.(4) Nevertheless, the removal of race correction in eGFR calculation has several ramifications for clinical decision making extending beyond the reviewed scenarios.

We found in our study that using race-corrected eGFR calculations for AKI risk stratification, in fact, reduces the accuracy in both logistic regression and machine learning models for Black patients by incorrectly classifying these patients at lower risk. Although the machine learning model had better calibration in Black patients than regression-based models, it also significantly underestimated their risk. The removal of race from the eGFR calculation resulted in improvements in the calibration of the risk models for both Black and non-Black patients, which supports such a change.

Although the removal of the race correction factor in eGFR calculation reduces the bias of the current AKI risk stratification models, an important observation remains that Black patients are at a higher risk for AKI overall. Allowing interaction terms between race and other predictors significantly reduces the underestimation of risk among Black patients, indicating the possibility of differential effects of risk factors on AKI between Black and non-Black patients. Race was not included in the list of candidate variables during the development of both the NCDR model and our ML models. The premise behind this decision, as is widely accepted, is that race is a social and not a biologic construct(12) and the inclusion of race in treatment decisions has the potential to perpetuate disparities in care delivery. However, we observe that the risk model with eGFR including the race correction has good calibration in the overall population and the under-estimation is only seen in Black patients, and the model calibration improves with inclusion of race variables. This calls for collection of richer data and a deeper dive into the data using novel approaches so that the uncaptured determinants of this elevated risk among Black patients whether biological or non-biological can be more accurately incorporated.

Further, although the calibration improves with the incorporation of interaction terms for race, the AUC values (discrimination) do not meaningfully change, in the overall population or in race groups. The AUC is also significantly lower in the Black group compared with non-Black group. This suggests that for further improvements to the AKI risk model there is a need to consider additional risk factors that may be differentially important across race groups.

The study has several limitations. First, the NCDR is not representative of PCI patients in non-US centers and the performance of risk models overall and the effect of race inclusion in eGFR calculation on AKI risk may differ for these populations. Second, the risk models were based on in-hospital AKI. Several patients did not have post procedure creatinine assessed and may have missed AKI and many patients may have developed AKI after discharge. Missing these patients and events can compromise the performance of the risk models.

## CONCLUSION

Removal of race correction in eGFR calculation from the contemporary post-PCI AKI risk models improves the accuracy of risk stratification for Black patients supporting its immediate implementation in line with the NKF and ASN task force recommendations. However, its removal does not eliminate the systematic underestimation of risks for Black patients. In light of this, there is a need to better capture the determinants of higher AKI risk among Black patients through richer data and advanced modeling techniques.

## Data Availability

Data are owned and were provided by the American College of Cardiology (ACC)’s National Cardiovascular Data Registry (NCDR), Washington, DC. Interested researchers can apply for data access by going to www.ncdr.com, clicking “Research”, and submitting a Research Proposal Application. Access to the CathPCI registry data will be granted when an agreement with ACC is reached.

## REFERENCES

1. Eneanya ND, Yang W, Reese PP. Reconsidering the Consequences of Using Race to Estimate Kidney Function. JAMA 2019;322:113–114.

2. Levey AS, Bosch JP, Lewis JB, Greene T, Rogers N, Roth D. A More Accurate Method To Estimate Glomerular Filtration Rate from Serum Creatinine: A New Prediction Equation. Ann Intern Med 1999;130:461–470.

3. Levey AS, Stevens LA, Schmid CH et al. A new equation to estimate glomerular filtration rate. Ann Intern Med 2009;150:604–612.

4. Delgado C, Baweja M, Crews DC et al. A Unifying Approach for GFR Estimation: Recommendations of the NKF-ASN Task Force on Reassessing the Inclusion of Race in Diagnosing Kidney Disease. American Journal of Kidney Diseases.

5. Inker LA, Eneanya ND, Coresh J et al. New Creatinine- and Cystatin C–Based Equations to Estimate GFR without Race. New England Journal of Medicine 2021;385:1737–1749.

6. Tsai TT, Patel UD, Chang TI et al. Validated contemporary risk model of acute kidney injury in patients undergoing percutaneous coronary interventions: insights from the National Cardiovascular Data Registry Cath-PCI Registry. J Am Heart Assoc 2014;3:e001380.

7. Gurm HS, Dixon SR, Smith DE et al. Renal Function-Based Contrast Dosing to Define Safe Limits of Radiographic Contrast Media in Patients Undergoing Percutaneous Coronary Interventions. Journal of the American College of Cardiology 2011;58:907–914.

8. Raposeiras-Roubín S, Abu-Assi E, Ocaranza-Sánchez R et al. Dosing of iodinated contrast volume: A new simple algorithm to stratify the risk of contrast-induced nephropathy in patients with acute coronary syndrome. Catheterization and Cardiovascular Interventions 2013;82:888–897.

9. Huang C, Murugiah K, Mahajan S et al. Enhancing the prediction of acute kidney injury risk after percutaneous coronary intervention using machine learning techniques: A retrospective cohort study. PLoS Med 2018;15:e1002703.

10. Mehta RL, Kellum JA, Shah SV et al. Acute Kidney Injury Network: report of an initiative to improve outcomes in acute kidney injury. Crit Care 2007;11:R31–R31.

11. Jones CA, McQuillan GM, Kusek JW et al. Serum creatinine levels in the US population: third National Health and Nutrition Examination Survey. Am J Kidney Dis 1998;32:992–9.

12. Fontanarosa PB, Bauchner H. Race, Ancestry, and Medical Research. Jama 2018;320:1539–1540.

