## Supplemental Tables for "Effect of removing race correction factor in glomerular filtration rate estimation on predicting acute kidney injury after percutaneous coronary intervention"

**Table S1. Performance comparison of logistic regression models in external validation set.**

|  | **Calibration** | | | | | | **Discrimination** | | |
| --- | --- | --- | --- | --- | --- | --- | --- | --- | --- |
|  | **Mean risk in overall population, %** | | **Mean risk in Black patients, %** | | **Mean risk in non-Black patients, %** | | **AUC in overall population** | **AUC in Black patients** | **AUC in non-Black patients** |
|  | **Observed** | **Predicted** | Observed | Predicted | Observed | Predicted |  |  |  |
| Model 1 | 7.5(7.4-7.5)^a^ | 7.4(7.4-7.4) | 10.4(10.3-10.5) | 7.7(7.7-7.8) | 7.2(7.2-7.2) | 7.4(7.4-7.4) | 0.74(0.74-  0.74) | 0.71(0.71-  0.72) | 0.74(0.74-0.74) |
| Model 2 | 7.5(7.4-7.5) | 7.4(7.4-7.4) | 10.4(10.3-10.5) | 8.3(8.2-8.3) | 7.2(7.2-7.2) | 7.3(7.3-7.4) | 0.74(0.74-  0.74) | 0.71(0.71-  0.72) | 0.74(0.74-0.74) |
| Model 3 | 7.5(7.4-7.5) | 7.4(7.4-7.4) | 10.4(10.3-10.5) | 9.4(9.3-9.4) | 7.2(7.2-7.2) | 7.2(7.2-7.3) | 0.74(0.74-  0.74) | 0.71(0.71-  0.72) | 0.74(0.74-0.74) |

^a^Results are shown in mean (95% confidence interval).

Model 1: Logistic regression model with race factor included in the eGFR calculation. Model 2: Model with race factor removed from eGFR calculation. Model 3: Model with interactions between race and age, prior cardiogenic shock, cardiac arrest, baseline eGFR, anemia, CAD presentation, diabetes and heart failure within 2 weeks.

AUC: area under the receiver operating characteristic curve; eGFR: estimated glomerular filtration rate; CAD: coronary artery disease.

**Table S2. Performance comparison of machine learning models in external validation set.**

|  | Calibration | | | | | | Discrimination | | |
| --- | --- | --- | --- | --- | --- | --- | --- | --- | --- |
|  | Mean risk in overall population, % | | Mean risk in Black patients, % | | Mean risk in non-Black patients, % | | AUC in overall population | AUC in Black patients | AUC in non-Black patients |
|  | Observed | Predicted | Observed | Predicted | Observed | Predicted |  |  |  |
| Model 1 | 7.5(7.4-7.5)^a^ | 7.4(7.4-7.4) | 10.4(10.3-10.5) | 8.3(8.3-8.4) | 7.2(7.2-7.2) | 7.3(7.3-7.3) | 0.77(0.77-0.77) | 0.74(0.73-0.74) | 0.78(0.77-0.78) |
| Model 2 | 7.5(7.4-7.5) | 7.4(7.4-7.4) | 10.4(10.3-10.5) | 8.6(8.6-8.6) | 7.2(7.2-7.2) | 7.3(7.3-7.3) | 0.77(0.77-0.77) | 0.74(0.74-0.75) | 0.78(0.77-0.78) |
| Model 3 | 7.5(7.4-7.5) | 7.4(7.4-7.4) | 10.4(10.3-10.5) | 9.4(9.3-9.4) | 7.2(7.2-7.2) | 7.2(7.2-7.2) | 0.77(0.77-0.78) | 0.74(0.74-0.75) | 0.78(0.77-0.78) |

^a^Results are shown in mean (95% confidence interval).

Model 1: Machine learning model with race factor included in the eGFR calculation. Model 2: Model with race factor removed from eGFR calculation. Model 3: Model with race as an independent predictor.

AUC: area under the receiver operating characteristic curve; eGFR: estimated glomerular filtration rate; CAD: coronary artery disease.
